# DETECTION OF ITALIAN NURSES’ THOUGHTS ON SEXUAL HEALTH AND THEIR KNOWLEDGE ON THE HIV-TRANSMISSION: A WEB SURVEY

**DOI:** 10.1101/2021.05.09.21251669

**Authors:** Italo Costanzo

**Affiliations:** Student at University of Molise, Department of Medicine and Health Sciences “Vincenzo Tiberio”, Via Francesco De Sanctis 1, 86100, Campobasso, Italy; Clinical Nurse Specialist at Integrated Contraception and Sexual Health Services, Cambridgeshire Community Services NHS Trust, Meadow Lane, Saint Ives, PE27 4LG, United Kingdom

**Author notes:** Corresponding author: Italo Costanzo.

**Keywords:** *Nursing Education* (MeSH Unique ID D004506), *HIV* (MeSH Unique ID D006678), *Surveys and Questionnaires* (MeSH Unique ID D011795), *Sexual Health* (MeSH Unique ID D000074384), *Italy* (MeSH Unique ID D007558)

## Abstract

**INTRODUCTION:** Despite the prevention of sexually transmitted infections included HIV and promotion of sexual health are fundamental parts of human health, the education system in Italy does not seem to provide healthcare professionals with specific knowledge requested by the most recent discoveries on HIV transmission.

**METHODS:** A web survey was randomly administered to Italian nurses to collect their thoughts about the academic education they received and analyse their knowledge on one of the newest discoveries related to HIV transmission.

**RESULTS:** More than half of the participants never received any specific academic training on sexual health and sexually transmitted infections, and more than three-quarters of the respondents did not know that a gay couple where only one of the partners is HIV positive will not have to use condoms forever and for every sexual intercourse.

**CONCLUSIONS:** The survey suggests that training on sexual health and sexually transmitted infections of Italian nurses need to be reinforced and it also highlights the need of updating nurses on the <undetectable equals untransmittable> campaign.

**KEY POINT:** Knowledge related to sexual health and HIV needs to be expanded in Italian nurses.

## BACKGROUND

Sexual health, defined as physical, mental and social wellbeing related to sexuality, is fundamental for its psychophysical, social, motivational and behavioral consequences on every person’s life[12]. Inadequate sexual health consequences represent an high social cost and an important public health problem that nurses have to deal with every time there is the opportunity[3-4].

Though sexual health is a specialist field of nursing, every nurse should be be part of the STI’s prevention, sexual health promotion and patients education at every level[5-7] and therefore, nursing education should be relevant and directed to the development of a positive attitude and communication ability[8-9].

This web survey was conducted with the aim to understand the role played by the university in providing nurses with sexual health knowledge and abilities and to investigate the thoughts of Italian nurses on the education received, the relationship between education and quality of care, and the level of knowledge related to the latest discovery on HIV transmission.

## METHODS

Ethical approval for this survey was obtained retrospectively on the 06/05/2021 from the ethical committee of the University of Molise with protocol number 17308.

Each participant gave their informed consent by voluntary and anonymously answering all the questions of the questionnaire.

The author was not aware of the identity of the participants and no participant has come or will become aware of the identity of the others.

An explanatory letter on the aim of the survey, risks and potential benefits resulting from the participation in the study was included in the link of the survey.

The survey was created with a professional software and was randomly spread in 2018/2019 on the main Italian nursing groups and blogs on social networks, allowing a simple diffusion in a wide geographical area and limitation of costs.

The questionnaire, composed by clear and explicit questions, was realized in collaboration with a panel of healthcare professionals and included three parts: four questions identifying age, sex, highest qualification obtained and current work environment; seven multiple-choice questions on received nursing education, clinical experience and nurses’ thoughts; one final question related to the latest discovery on HIV transmission.

Failing to reply to just one of the questions did not allow the transition to the next question, therefore the totality of the data required was obtained for each participant.

## RESULTS

105 nurses participated in the survey, most of them aged 20-29 (tab. 1), female (tab. 2), holding a BSc Nursing degree or equivalent (tab. 3) and working in a medical or surgical area (tab. 4).

**Table 1.** How old are you?

| AGE | FREQUENCY | PERCENTAGE |
| --- | --- | --- |
| 20-24 | 30 | 28.57 |
| 25-29 | 37 | 35.24 |
| 30-34 | 9 | 8.57 |
| 35-39 | 6 | 5.71 |
| 40-44 | 6 | 5.71 |
| 45-49 | 10 | 9.52 |
| 50 or over | 7 | 6.67 |
| TOTAL | 105 | 100.00 |

**Table 2.** What is your gender?

| GENDER | FREQUENCY | PERCENTAGE |
| --- | --- | --- |
| Male | 25 | 23.81 |
| Female | 80 | 76.19 |
| Prefer not to say | 0 | 0.00 |
| TOTAL | 105 | 100.00 |

**Table 3.** What is the highest qualification you obtained?

| QUALIFICATION | FREQUENCY | PERCENTAGE |
| --- | --- | --- |
| Nursing student | 10 | 9.52 |
| BSc Nursing or equivalent | 59 | 56.19 |
| MSc Nursing or equivalent | 36 | 34.29 |
| PhD or equivalent | 0 | 0.00 |
| TOTAL | 105 | 100.00 |

**Table 4.** What is your current area of work/practice?

| CURRENT WORK AREA | FREQUENCY | PERCENTAGE |
| --- | --- | --- |
| Medical or surgical area | 37 | 35.24 |
| Technical area | 13 | 12.38 |
| Critical/emergency area | 33 | 31.43 |
| Specialist area | 22 | 20.95 |
| TOTAL | 105 | 100.00 |

As shown in fig. 1, 58.10% of nurses (61 units) did not receive specific training on sexual health and STI’s during their education, unlike the remaining 41.90% (44 units).

**Figure 1.**
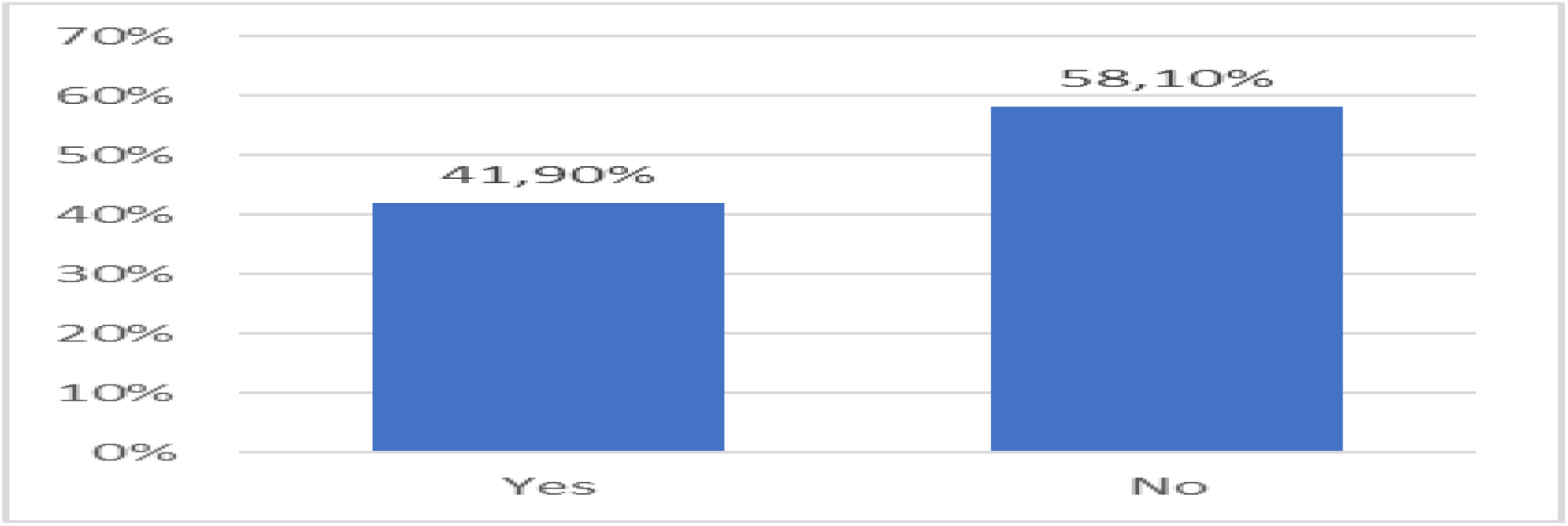
Education - Have you received any specific training on sexual health and STI’s during your training?

As shown in fig. 2, 70.48% (74 units) of the participants believe that gaining more knowledge in sexual health and STI’s would improve their clinical practice, while 10.48% (11 units) believe that it would not and 19.05% (20 units) state to be unable to answer the question.

**Figure 2.**
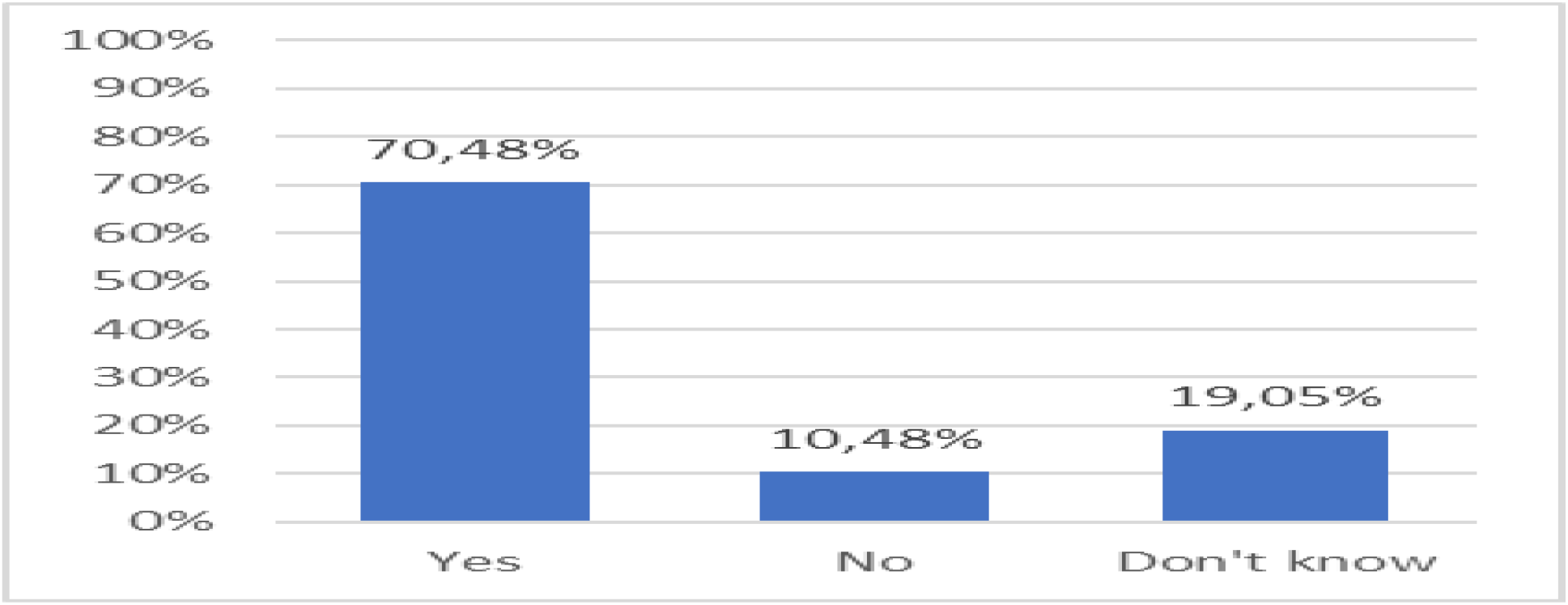
Relationship between knowledge gain and clinical practice – Do you think that gaining more knowledge on sexual health and STI’s would improve your clinical practice?

Fig. 3 shows that 53.33% (56 units) of nurses faced issues, problems or questions related to sexual health coming from a patient while they were caring for them at least once, while for 46, 67% (49 units) it never happened.

**Figure 3.**
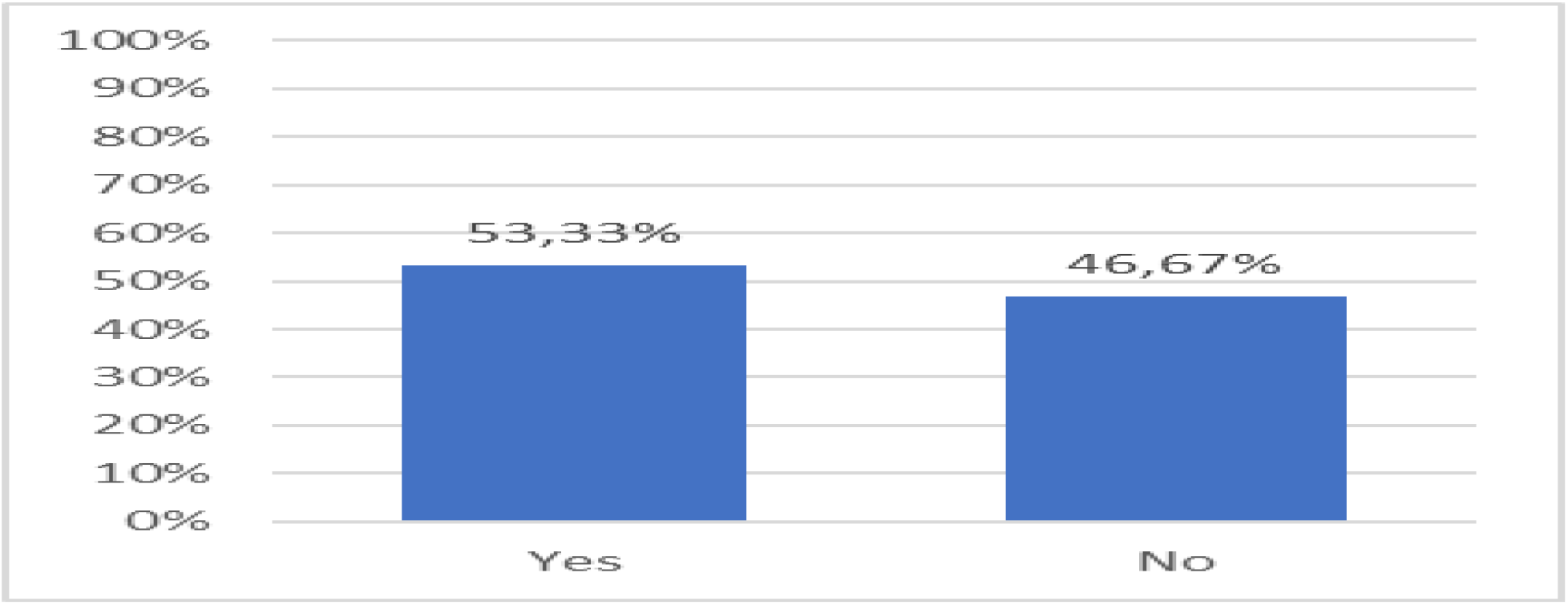
Clinical Practice - Have you ever faced issues, problems or questions related to sexual health coming from a patient you were caring for?

When dealing with sexual health issues, 76.19% (80 units) of nurses would request a specialist advice, unlike the remaining 23.81% (25 units) of the participants, as shown in fig. 4.

**Figure 4.**
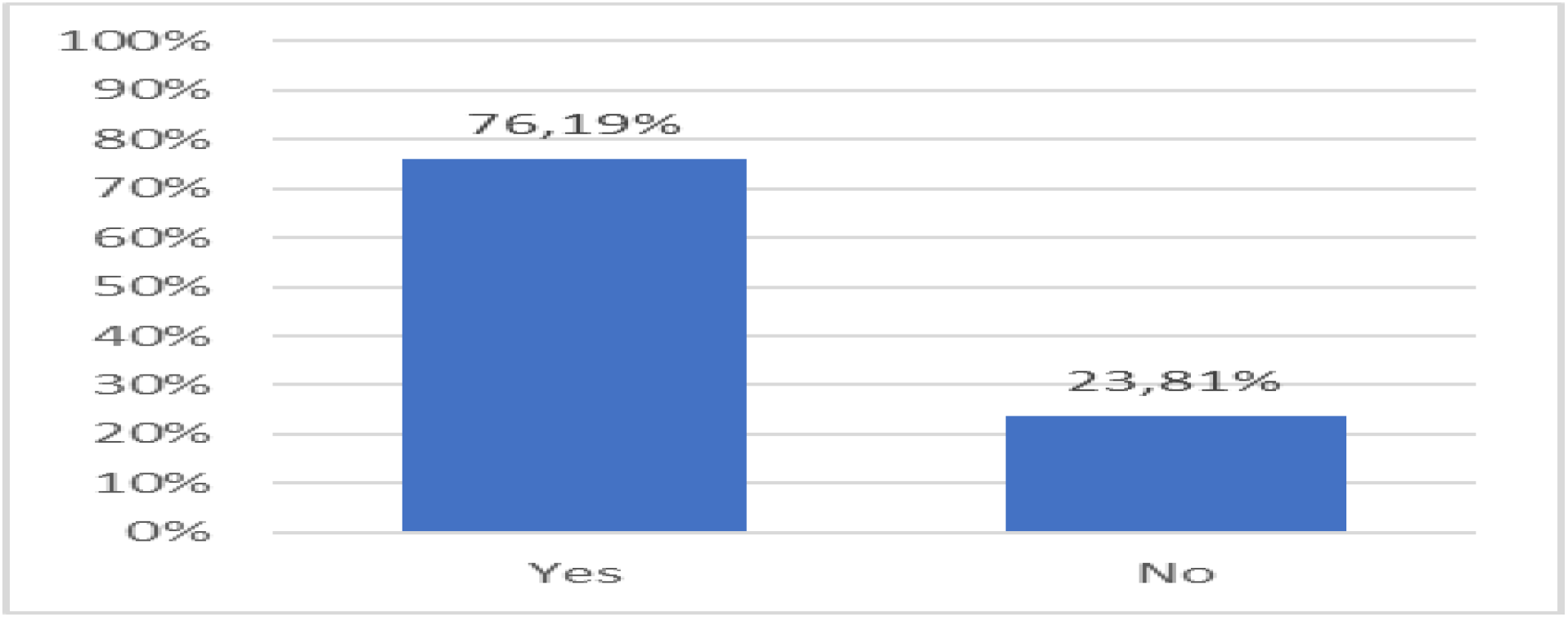
Consultation - If you were to care for a patient with sexual health needs, would you request a specialist advice?

Fig. 5 shows which of the healthcare professionals would probably be asked for help by the nurse when dealing with a sexual health matter: 37.14% (39 units) answered D “andrologist, urologist or gynecologist”, 33.33% (35 units) B “sexologist”, 33.33% E “psychologist or psychotherapist”, 30.48% (32 units) A “nurse or ward manager”, 10.48% (11 units) F “psychiatrist”, 5.71% (6 units) G “general practitioner” and 5.71% (6 units) C “others” (6 units).

**Figure 5.**
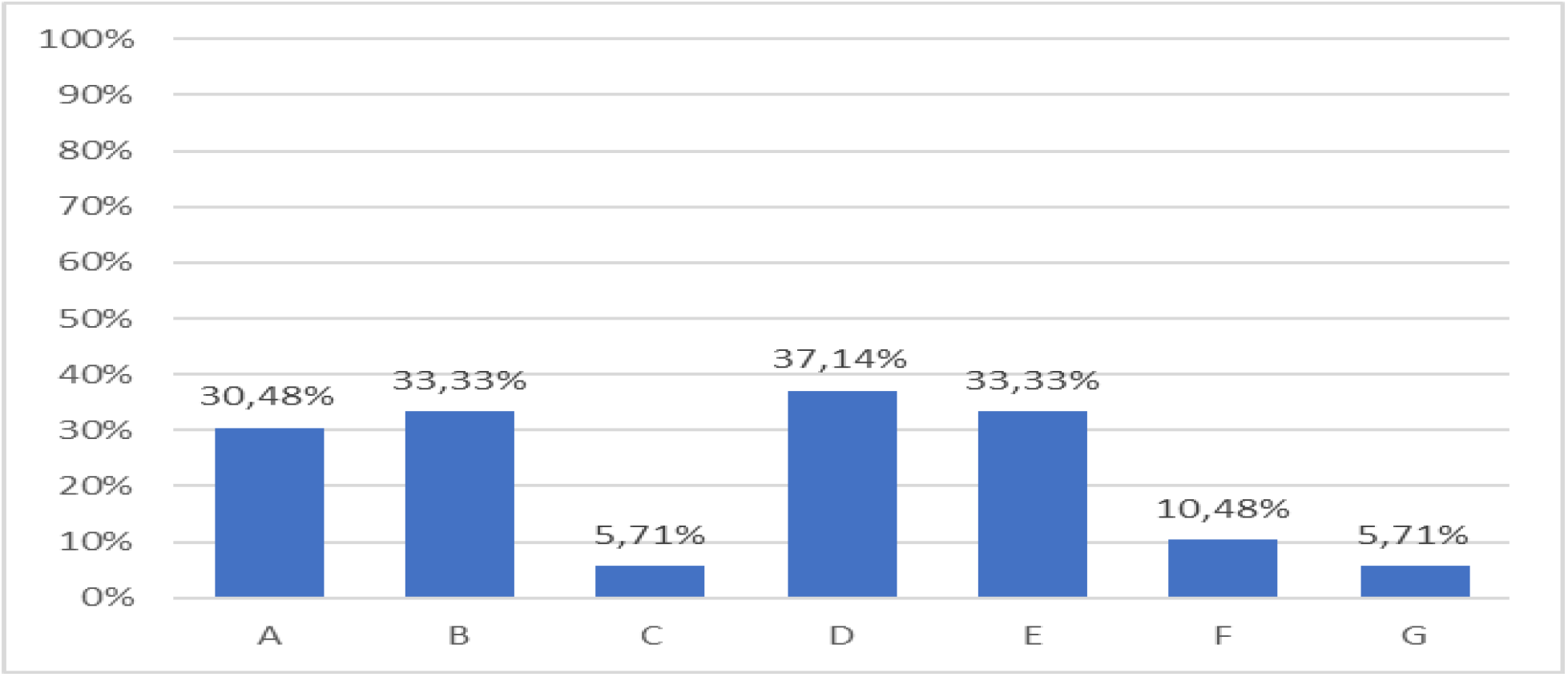
Healthcare professionals - If you need advice on issues related to a patient’s sexual health, which healthcare professional would you ask for help?

Fig. 6 shows that 50.48% of nurses (53 units) believe that sexual health and STI’s education has “enough” influence on the quality of nursing care provided to patients.

**Figure 6.**
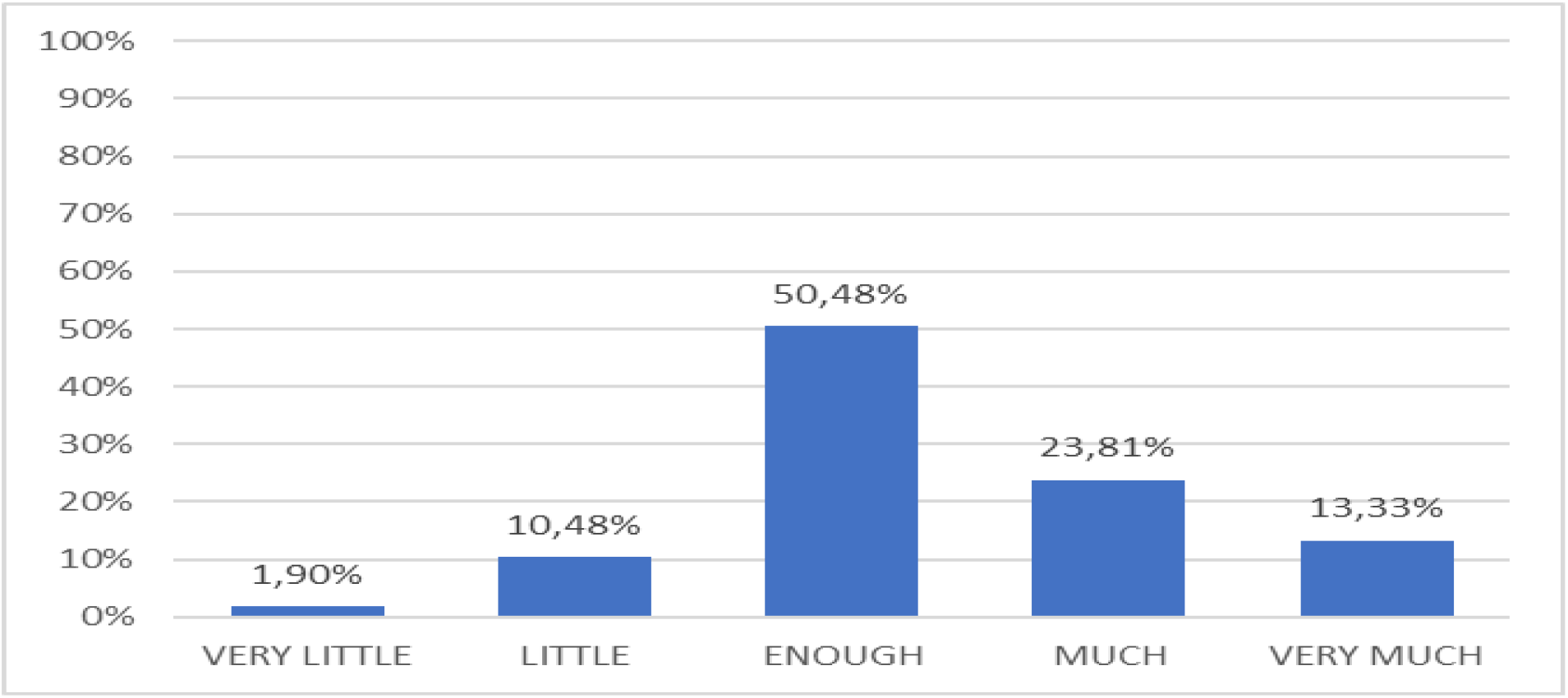
Relationship between education and quality of care - How much do you think that training on sexual health and STI’s affects the quality of nursing care provided to patients?

23.81% (25 units) answered “much”, 13.33% (14 units) answered “very much”, 10.48% (11 units) answered “little” and only 1.90% (2 units) responds “very little”.

Fig. 7 shows that 37.14% (39 units) of participants recognize reading books, magazines, newspapers and scientific articles as the main source of their knowledge (E), while 30.48% (32 units) recognize it in their nursing education (A). For 9.52% (10 units) the knowledge comes directly from clinical practice (B), for another 9.52% (10 units) from personal life experiences (F), for 7.62% (8 units) from watching TV programs, movies or documentaries (D) and for only 5.71% (6 units) from update courses (C).

**Figure 7.**
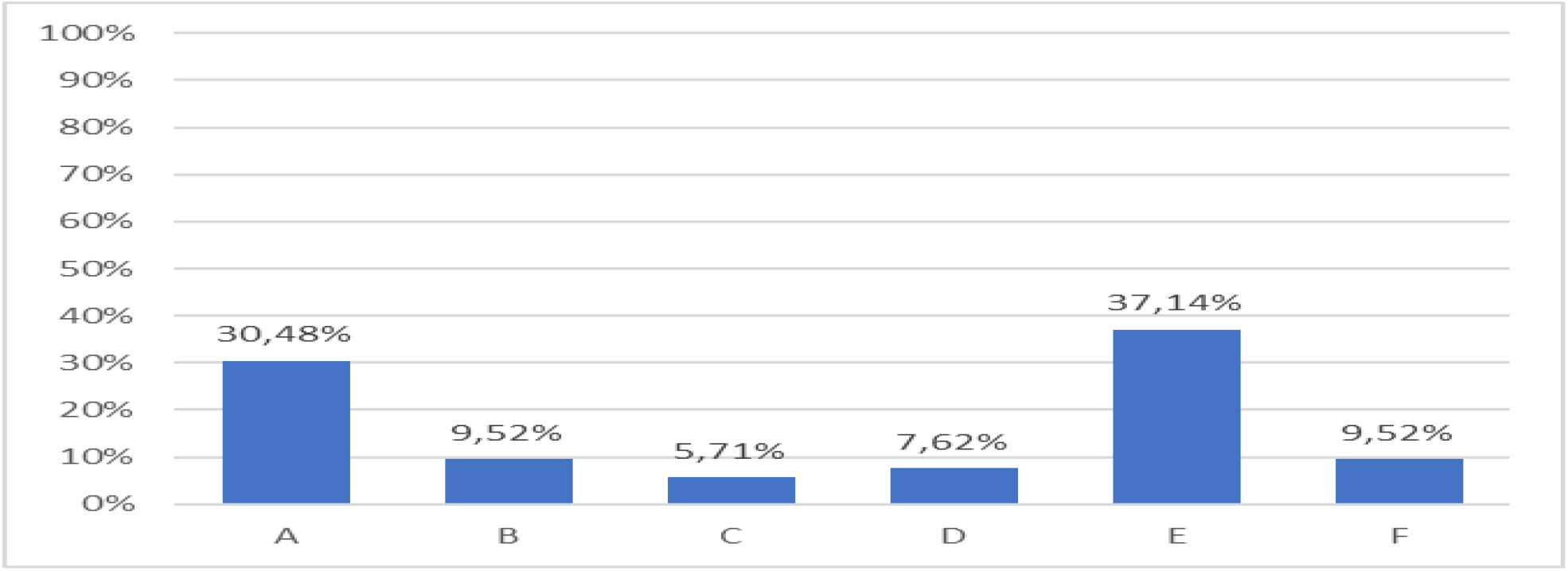
Main source of knowledge - Where do you think your knowledge about sexual health and STI’s comes from most?

**Table 1.** Quiz question: Should a gay couple in which one of the partners is HIV positive use the condom forever and for every sexual intercourse?

| YES | NO | CORRECT ANSWER | AVERAGE SCORE |
| --- | --- | --- | --- |
| 91 nurses (86,67%) | 14 nurses (13,33%) | NO | 13% |

The fact that only 13% of participants answered correctly shows that most of the nurses were not updated to the latest research on HIV transmission, stating that the risk of HIV transmission in gay couples through condom-less sex when HIV viral load is suppressed is effectively zer o and supporting the message of the “U=U” (undetectable equals untransmittable) campaign[10].

## DISCUSSION

Most Italian nurses never received any specific training on sexual health and STI’s and acquired their sexual health-related knowledge outside of the university, mainly by reading independently.

Because Italian nurses recognize the negative impact that lack of sexual health-related education has on clinical practice and quality of care and most nurses do come across with patients experiencing sexual health issues, about 70% of participants believe that more specific knowledge gain on sexual health would improve their clinical practice.

The role of the nurse is essential in maintaining people’s sexual health at every stage of life and although the reluctance to manage this type of human needs may origin from embarrassment, lack of time, fear of offending, conservative opinions, not giving the problem the right priority or believing that patients do not expect them to nurses face these problems with them, the literature well reports this often happens because nurses are afraid of not being able to respond effectively to issues raised by patients due to lack of knowledge or experience[11-20].

It seems that Italian nurses know that sexual health is a fundamental part of patients’ wellbeing and that nurses attitude towards sexuality can affect patients care[21-24], in fact more than threequarters of the participants believe that sexual health and STI’s-related education has an impact on the quality of nursing care provided.

Even if most of the nurses would consult with a specialist when dealing with patients’ sexual health matters, there is certainly no clear and unambiguous perception as to which professional they can turn to and despite the current growing autonomy and professionalization of the nurses in Italy, only 30% of the participants would ask another nurse or the ward manager for help with sexual health-related issues. The healthcare professionals they considered most suitable are the urologist, the andrologist and the gynaecologist (37%), followed by the sexologist, the psychologist and the psychotherapist (33%).

The average score for the final question is 13%, therefore it clearly demonstrates that Italian nurses do need an update on the most recent research on HIV transmission.

## LIMITS

Although some difficulty was encountered with the enrolment of a statistically significant sample, the survey sheds light on something that was never previously analysed and offers basic data to better understand the educational needs of Italian nurses and improve the quality of care in the Italian national health service.

## CONCLUSIONS

Specialist knowledge in sexual health, sexually transmitted infections and HIV of Italian nurses needs to be expanded and strengthened. A nursing education review is therefore suggested to make sure sexual health is included in every nursing training program.

## Data Availability

No extra data available.

